# Healthcare utilization among adults with co-occurring substance use and mental health disorders (2018-2023): A study based on All of Us program

**DOI:** 10.64898/2026.01.28.26344935

**Authors:** Abdul-Hanan Saani Inusah, Muyang Wu, Zoey Babyak, Xiaoming Li, Shan Qiao

## Abstract

**Background:** Co-occurring substance use and mental health disorders (COD) represent a growing public health concern, yet healthcare utilization studies with a large sample size remain limited. This study examined healthcare utilization patterns and sociodemographic correlates among COD adults using data from the All of Us Research Program (2018–2023).

**Methods:** Electronic health record data were analyzed for adults aged ≥18 years with confirmed diagnoses of substance use and mental health disorders recorded on at least two occasions. Healthcare services were identified using the standardized Current Procedural Terminology and Healthcare Common Procedure Coding System codes and categorized into counseling and therapy, medication/somatic services, online or telehealth care, and other supportive modalities. Multivariable logistic regression was employed to assess sociodemographic and structural correlates of healthcare utilization.

**Results:** Among 19,423 adults with COD, 57.1% received healthcare. Counseling and therapy accounted for the largest share of encounters, while online services surged in 2020 during the COVID-19 pandemic. Healthcare utilization was higher among older adults (≥65 years: aOR=1.52, 95%CI:1.29–1.78), males (aOR=1.19, 95%CI:1.12–1.26), individuals with disabilities (aOR=1.46, 95%CI:1.36–1.56), and those with employer-sponsored (aOR=1.22, 95%CI:1.10–1.36) or other private insurance (aOR=2.15, 95%CI:1.97–2.34). The level of healthcare utilization was lower among participants with lower income (≤$25,000: aOR=0.75, 95%CI:0.69–0.81) or Medicaid coverage (aOR=0.83, 95%CI:0.77–0.89).

**Conclusions:** Despite high clinical need, healthcare utilization among adults with COD remains suboptimal and is shaped by structural inequities across income and insurance lines. Findings highlight the need to expand integrated healthcare services, strengthen Medicaid coverage, and sustain telehealth infrastructure to promote equitable, long-term engagement in care.

**Highlights:**

º Individuals with co-occurring disorders continue to face low healthcare utilization.
º Counseling and therapy were the major mode of care, while telehealth peaked during COVID-19.
º Lower income and Medicaid coverage were tied to lower healthcare utilization.
º Older adults and people with disabilities were more likely to use healthcare services.
º Findings highlight the needs to expand integrated, equitable behavioral care.

## Introduction

Co-occurring substance use and mental health disorders (COD) affect more than 21 million adults in the United States (U.S.), reflecting the high overlap between substance use disorders (SUD) and mental health disorders (MHD) (Substance Abuse and Mental Health Services Administration, 2022). Individuals with COD face elevated morbidity, premature mortality, and profound social and functional impairment compared with those who have a single disorder (Substance Abuse Mental Health Services Administration, 2021). These patterns emphasize a critical need to understand how individuals with COD engage with formal healthcare systems.

Despite substantial clinical need, healthcare utilization remains strikingly low (Substance Abuse and Mental Health Services Administration, 2022). Some studies indicate that fewer than one in ten adults with COD receive specialty substance use healthcare, and only a small fraction engage in integrated services addressing both conditions (Flynn & Brown, 2008; Priester et al., 2016). Most adults with COD either receive care for only one disorder, typically mental health, or receive no care at all. This pattern reflects long-standing fragmentation between addiction and mental health service systems (Substance Abuse Mental Health Services Administration, 2021).

Besides the low rate of healthcare utilization, access to healthcare is strongly shaped by structural and sociodemographic factors. Individuals with lower income or without insurance are consistently less likely to receive care for mental health or substance use conditions, even when their clinical needs are comparable to those of more advantaged peers (Dhinsa et al., 2023; Priester et al., 2016). Racial and ethnic minority groups also face disproportionate barriers to both types of care, with persistent differences in diagnosis, initiation and adequacy of treatment across multiple U.S. studies (Acevedo et al., 2018). However, limited evidence exists on how these determinants operate specifically within COD populations, where multiple intersecting barriers, economic, racial, and systemic, may interact to reduce healthcare utilization.

Most prior research on COD healthcare utilization has been cross-sectional or confined to specific diagnostic pairs (e.g., depression and alcohol use disorder), leaving the broader COD population understudied. Some longitudinal work has followed particular combinations of disorders with a relatively narrow scope (Lu et al., 2021; Worley et al., 2012). Few studies have leveraged large, nationally diverse electronic health record (EHR) datasets capable of capturing real-world utilization patterns over time. Moreover, existing research seldom examines how sociodemographic and structural correlates shape these utilization trajectories, including variations by insurance type, income, and disability status. Addressing this knowledge gap is critical to designing integrated behavioral health systems responsive to diverse patient needs.

In this study, we used EHR data from the nationally diverse All of Us Research Program to examine healthcare utilization among adults with co-occurring SUD and MHD between 2018 and 2023. We assessed the proportion of individuals who engaged in any documented SUD or MHD healthcare, defined using standardized Current Procedural Terminology (CPT), Healthcare Common Procedure Coding System (HCPCS), and Systematized Nomenclature of Medicine—Clinical Terms (SNOMED-CT) codes and described patterns across major service categories (counseling and therapy, medication/somatic care, online or telehealth, and other intensive or supportive services). We further evaluated how key sociodemographic and structural factors, including insurance coverage, income, employment, age, sex, race/ethnicity, and disability, were associated with healthcare utilization. Using a large, contemporary EHR dataset, this study provides new empirical evidence on the sociodemographic and structural correlates of healthcare utilization among adults with COD and identifies subgroups at greatest risk of underutilization.

## Methods

### Data Source

We used data from the All of Us Research Program, a nationwide precision medicine initiative launched by the National Institutes of Health (NIH) in 2018 with the goal of recruiting at least one million U.S. adults. Participants contribute a broad range of data, including EHR, survey responses, biospecimens, physical measurements, and wearable data. All data are de-identified and standardized using the Observational Medical Outcomes Partnership (OMOP) Common Data Model (CDM) and made available to approved researchers through a secure, cloud-based Researcher Workbench (All of Us Research Program Investigators, 2019; National Institutes of Health, 2025; Ramirez et al., 2022).

As of the most recent data release, the All of Us Research Program includes over 633,000 participants. For this study, healthcare-related encounters were identified from the Procedure Occurrence table, which records diagnostic and therapeutic services using standardized CPT, HCPCS, and SNOMED CT vocabularies (All of Us Research Program, 2025).

### Study Design and Cohort Construction

We conducted a retrospective observational analysis of EHRdata from January 1, 2018, through December 31, 2023. Adults aged 18 years or older with documented SUD and MHD were identified using International Classification of Diseases, Tenth Revision, Clinical Modification (ICD-10-CM) codes. To improve diagnostic certainty, each disorder was evaluated independently. Participants were classified as having co-occurring disorders (COD) if their EHR contained at least two encounters on different dates for SUD and at least two encounters on different dates for MHD during the study period. The two conditions were not required to be recorded during the same encounter, and this definition does not allow determination of whether SUD and MHD occurred concurrently or the temporal order in which they developed. Requiring multiple encounters for each disorder reduced potential misclassification arising from single-encounter or provisional codes that may reflect screening or suspected rather than confirmed diagnoses. SUD was defined using ICD-10-CM codes F10–F16 and F18–F19, and MHD using codes F20–F91, including all primary and sub-codes (*Supplementary Table 1*). A total of 27,494 participants met criteria for confirmed SUD. Of these, 19,423 also met confirmation criteria for MHD, and therefore constituted the analytic cohort of adults with COD for this study. The cohort selection process and inclusion flow are summarized in Figure 1.

**Figure 1.**
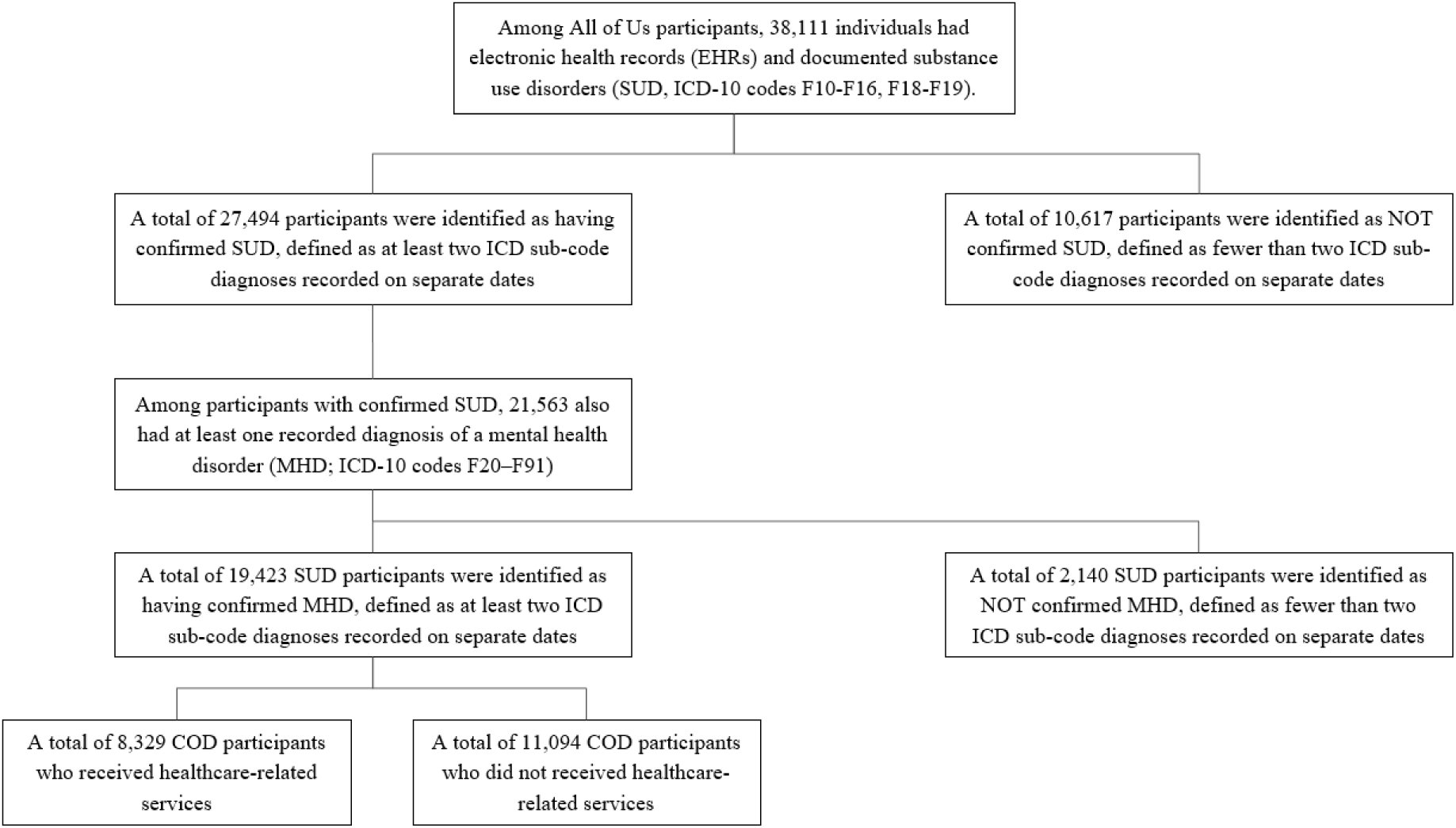
Selection of participants with confirmed co-occurring substance use and mental health disorders (COD) and healthcare utilization in the All of Us Research Program, 2018–2023. The flowchart illustrates the stepwise inclusion of adults with confirmed substance use disorders (SUD) and mental health disorders (MHD) based on ICD-10-CM diagnostic criteria. Participants were required to have at least two encounters with distinct diagnostic codes on separate dates for both SUD (F10–F16, F18–F19) and MHD (F20–F91). The final analytic cohort (n = 19,423) was divided into those who utilized healthcare services and those who did not.

### Healthcare Service Code Identification

To identify healthcare services in the EHR, we referenced multiple authoritative sources, including SimiTree’s listing of commonly billed CPT codes for substance use care, MediBillMD’s 2024 compendium of rehabilitation codes, and national HCPCS H-code directories (AAPC, 2025; Barnes, 2024; HCPCS Codes, 2025; SimiTree, 2024). Additional guidance was drawn from state Medicaid manuals such as Wisconsin Forward Health (Wisconsin Department of Health Services, 2025), the Centers for Medicare & Medicaid Services (CMS) physician self-referral CPT/HCPCS list (Centers for Medicare & Medicaid Services, 2025), the American Academy of Family Physicians (AAFP) (Loskutova et al., 2020), and SAMHSA’s SBIRT coding guide (Substance Abuse and Mental Health Services Administration, 2024a). These resources document codes that capture a broad spectrum of healthcare modalities, including counseling, therapy, medication management and other medical services, rehabilitation, and online care, for individuals with substance use and mental health conditions. The complete list of CPT and HCPCS healthcare codes used in this study is presented in *Supplementary Table 2*.

## Measures

### Outcome

The primary outcome was receipt of any MHD or SUD-related healthcare service, defined as at least one CPT- or HCPCS-coded encounter for SUD or MHD healthcare during the study period (yes/no).

### Healthcare Service Categories

Healthcare encounters were grouped into four categories: (1) *counseling and therapy*, including psychotherapy, counseling, and case management; (2) *medication and somatic services*, including medication management and prescriber visits; (3) *online or telehealth healthcare*, including virtual counseling or therapy sessions; and (4) *other healthcare services*, including crisis intervention and acute care, detoxification services, intensive outpatient programs, residential healthcare, rehabilitation and supportive services, and specialized or other healthcare modalities.

### Sociodemographic and Structural Variables

Variables were derived from EHR and participant-reported survey data and categorized into age groups (18-29, 30-39, 40-49, 50-64, ≥ 65 years), sex assigned at birth, race (Asian/Other/Unknown, Black or African American, White), ethnicity (Hispanic/Latino, Not Hispanic/Latino), educational attainment (high school degree or more, less than high school degree), household income (≤$25,000, >$25,000), employment status, health insurance coverage (insured vs. uninsured), type of insurance (Medicaid, Medicare, employer/union plans, other health plans), and disability status (yes/no). Disability was defined broadly to include physical or mental impairments that limited mobility, self-care, cognitive functioning, or the ability to complete daily errands independently.

Several sociodemographic variables included an “unknown” category. In general, participants with missing, skipped, or “prefer not to answer” data were classified as “unknown” and retained in the analyses to preserve sample size. Participants whose sex at birth was recorded as “Skip,” “Prefer Not To Answer,” “Don’t Know,” “Not male, not female, prefer not to answer, or skipped,” or “No matching concept” were recoded as “Unknown.” Thus, “Unknown” indicates that the participant did not report sex at birth or reported a response other than male or female.

### Statistical Analysis

We summarized sociodemographic characteristics for COD participants overall and by healthcare service utilization status. Annual healthcare utilization trends were summarized as the proportion of unique COD participants with at least one healthcare encounter in each calendar year from 2018–2023. To account for variation in follow-up time and potential clustering of encounters within participants, descriptive analyses were restricted to unique participants per year. Multivariable logistic regression was used to assess sociodemographic and structural correlates of healthcare service utilization among COD participants. The multivariable model adjusted for age, sex assigned at birth, race/ethnicity, sexual orientation, educational attainment, household income, insurance status, employment status, and disability. All covariates were selected a priori based on theoretical relevance and prior literature on behavioral health service disparities.

Adjusted odds ratios (aORs) and 95% confidence intervals (CIs) were estimated, with statistical significance defined as p < .05. Analyses were conducted using R within the secure All of Us Researcher Workbench.

## Results

### Sample Characteristics

*Table 1* summarizes the demographic and socioeconomic characteristics of the analytic sample. Among the 19,423 adults with COD, 57.1% received healthcare. The cohort was predominantly middle-aged or older, with 61.2 % aged ≥ 50 years. Nearly half of the participants identified as White (49.4%), followed by Black or African American (24.3%). Most participants (80.8%) identified as non-Hispanic, while 15.2% identified as Hispanic or Latino. Educational attainment was high, with 92.6% reporting a high-school degree or higher. Half (50.0%) reported annual household incomes of ≤$25,000, and 71.4% were unemployed. Health-insurance coverage was reported by 89.5% of participants; 44.5% were enrolled in Medicaid, 21.5% in Medicare, 16.2% in other health plans, and 12.1% in employer- or union-based insurance. Disability was reported by 27.3% of participants. Statistically significant differences (p < 0.001) were observed across most sociodemographic variables between those who utilized healthcare service and those who did not.

**Table 1.** Sociodemographic and structural characteristics of adults with co-occurring substance use and mental health disorders (COD), by healthcare utilization (N = 19,423).

| Variable | Healthcare Utilization |  | Total<br>(N=19423) | P-Value |
| --- | --- | --- | --- | --- |
|  | Yes<br>(N=11094) | No<br>(N=8329) |  |  |
| **Age Group** |  |  |  | <0.001 |
| 18-29 | 538 (4.8%) | 339 (4.1%) | 877 (4.5%) |  |
| 30-39 | 1890 (17.0%) | 1137 (13.6%) | 3027 (15.6%) |  |
| 40-49 | 2161 (19.5%) | 1467 (17.6%) | 3627 (18.7%) |  |
| 50-64 | 4406 (39.7%) | 3252 (39.0%) | 7658 (39.4%) |  |
| 65+ | 2099 (18.9%) | 2134 (25.6%) | 4233 (21.8%) |  |
| **Race Group** |  |  |  | <0.001 |
| Asian/Other/Unknown | 3074 (27.7%) | 2023 (24.3%) | 5097 (26.2%) |  |
| Black or African American | 2791 (25.1%) | 1936 (23.3%) | 4727 (24.3%) |  |
| White | 5229 (47.1%) | 4370 (52.4%) | 9599 (49.4%) |  |
| **Ethnicity Group** |  |  |  | <0.001 |
| Hispanic or Latino | 1812 (16.3%) | 1145 (13.7%) | 2957 (15.2%) |  |
| Not Hispanic or Latino | 8817 (79.5%) | 6884 (82.7%) | 15701 (80.8%) |  |
| Unknown | 465 (4.2%) | 300 (3.6%) | 765 (3.9%) |  |
| **Sex** |  |  |  | <0.001 |
| Female | 5426 (48.9%) | 3551 (42.6%) | 8977 (46.2%) |  |
| Male | 5397 (48.6%) | 4543 (54.5%) | 9940 (51.2%) |  |
| Unknown | 271 (2.4%) | 235 (2.8%) | 506 (2.6%) |  |
| **Sexual Orientation** |  |  |  | 0.0027 |
| Heterosexual | 9171 (82.7%) | 6746 (81.0%) | 15917 (81.9%) |  |
| Non-Heterosexual | 1501 (13.5%) | 1275 (15.3%) | 2776 (14.3%) |  |
| Unknown | 422 (3.8%) | 308 (3.7%) | 730 (3.8%) |  |
| **Education** |  |  |  | 0.0016 |
| High school degree or more | 10204 (92.0%) | 7775 (93.3%) | 17979 (92.6%) |  |
| Less than a high school degree | 454 (4.1%) | 281 (3.4%) | 735 (3.8%) |  |
| Unknown | 436 (3.9%) | 273 (3.3%) | 709 (3.6%) |  |
| **Income** |  |  |  | <0.001 |
| > 25,000 US dollars | 2508 (22.6%) | 2873 (34.5%) | 5381 (27.7%) |  |
| ≤ 25,000 US dollars | 5893 (53.1%) | 3827 (45.9%) | 9720 (50.0%) |  |
|  | Yes<br>(N=11094) | No<br>(N=8329) |  |  |
| Unknown | 2692 (24.3%) | 1629 (19.6%) | 4321 (22.2%) |  |
| **Employment Status** |  |  |  | <0.001 |
| Employed for wages/self-employed | 2481 (22.4%) | 2155 (25.9%) | 4636 (23.9%) |  |
| Not employed | 8030 (72.4%) | 5831 (70.1%) | 13861 (71.4%) |  |
| Unknown | 583 (5.3%) | 343 (4.1%) | 926 (4.8%) |  |
| **Health Insurance** |  |  |  | <0.001 |
| Health Insurance: No | 708 (6.4%) | 668 (8.0%) | 1376 (7.1%) |  |
| Health Insurance: Yes | 9978 (90.0%) | 7403 (88.9%) | 17381 (89.5%) |  |
| Unknown | 408 (3.7%) | 258 (3.1%) | 666 (3.4%) |  |
| **Employer Or Union** |  |  |  | <0.001 |
| No | 9956 (89.7%) | 7110 (85.4%) | 17066 (87.9%) |  |
| Yes | 1138 (10.2%) | 1219 (14.6%) | 2357 (12.1%) |  |
| **Medicaid** |  |  |  | <0.001 |
| No | 5575 (50.3%) | 5210 (62.6%) | 10785 (55.5%) |  |
| Yes | 5519 (49.7%) | 3119 (37.4%) | 8638 (44.5%) |  |
| **Medicare** |  |  |  | <0.001 |
| No | 8818 (79.5%) | 6422 (77.1%) | 15240 (78.5%) |  |
| Yes | 2276 (20.5%) | 1907 (22.9%) | 4183 (21.5%) |  |
| **Other health plans** |  |  |  |  |
| No | 9939 (89.6%) | 6338 (76.1%) | 16277 (83.8%) |  |
| Yes | 1155 (10.4%) | 1991 (23.9%) | 3146 (16.2%) |  |
| **Disability** |  |  |  | <0.001 |
| No | 8484 (76.5%) | 5634 (67.6%) | 14118 (72.7%) |  |
| Yes | 2610 (23.5%) | 2695 (32.4%) | 5305 (27.3%) |  |

Notes: Values represent frequencies and percentages within each healthcare category. Statistical significance for group differences was assessed using chi-square tests.

### Healthcare Utilization Trends

From 2018 through 2023, counseling and therapy accounted for the largest share of healthcare utilization, with 2,149 to 3,321 participants receiving care and 25,547 to 54,386 total encounters recorded annually. Utilization peaked in 2019 and declined steadily thereafter. Medication and somatic services showed stable engagement, involving 1,871 to 1,970 participants per year and 16,641 to 27,312 encounters, with minimal year-to-year variation. Online healthcare demonstrated the most notable fluctuation, rising sharply in 2020 (1,636 patients; 10,420 encounters), coinciding with the COVID-19 telehealth expansion, before gradually decreasing in subsequent years. The “Other” category, comprising crisis intervention, detoxification, intensive outpatient, residential, and supportive services, remained consistently low, with fewer than 205 patients and 1,184 encounters per year. Overall, these trends highlight the centrality of counseling and therapy, the stability of medication-based care, and a transient surge in telehealth-driven healthcare during the pandemic (*Figures 2* and *3*).

**Figure 2.**
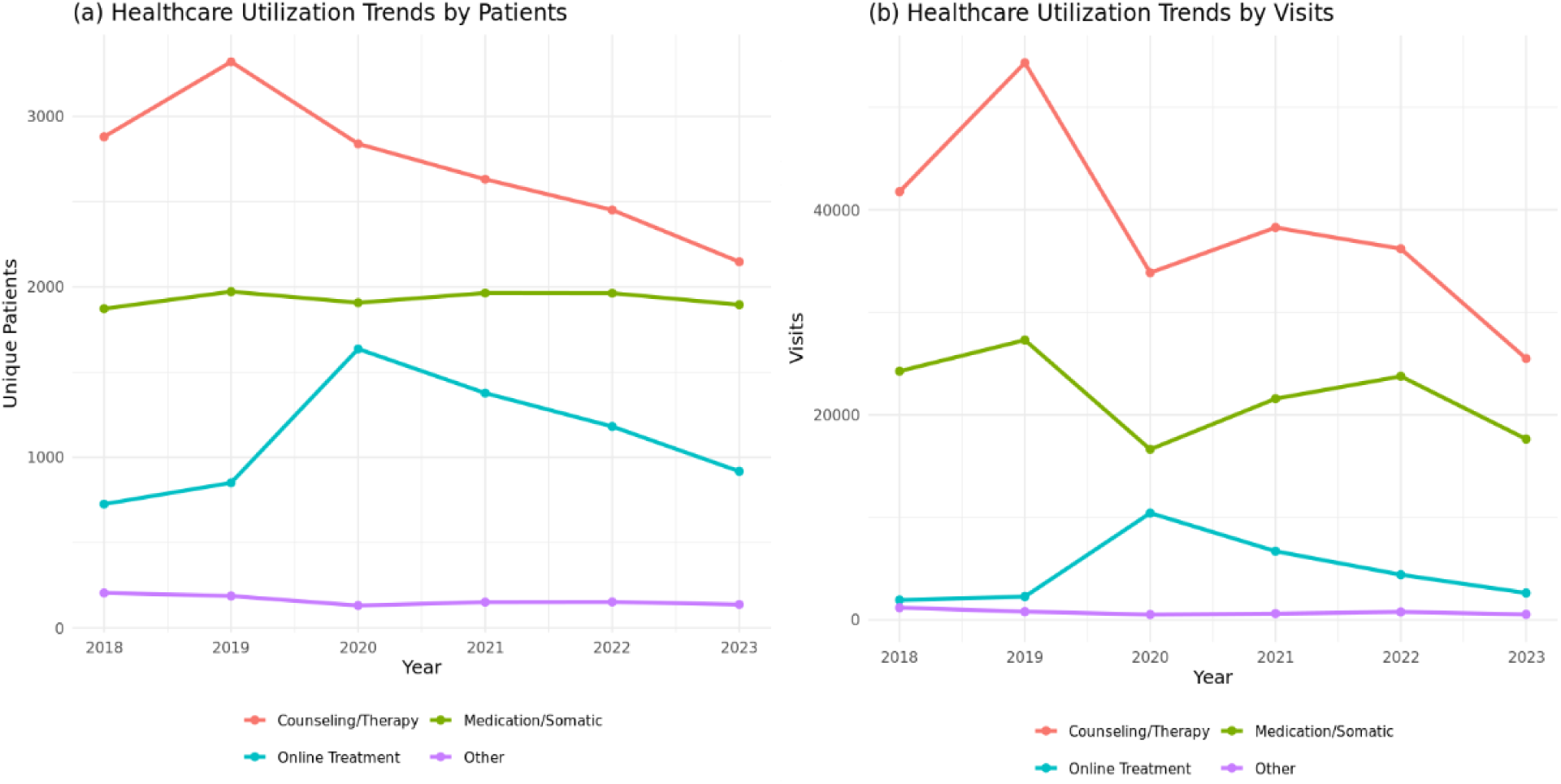
Healthcare utilization trends among adults with co-occurring substance use and mental health disorders (COD), All of Us Research Program, 2018–2023. *(a) Trends by unique patients* *(b) Trends by visits*. Counseling and therapy consistently represented the largest share of treatment utilization, while medication/somatic services remained relatively stable across years. Online healthcare peaked in 2020 during the COVID-19 pandemic before declining. The “Other” category represented the lowest-utilized services throughout the study period.

**Figure 3.**
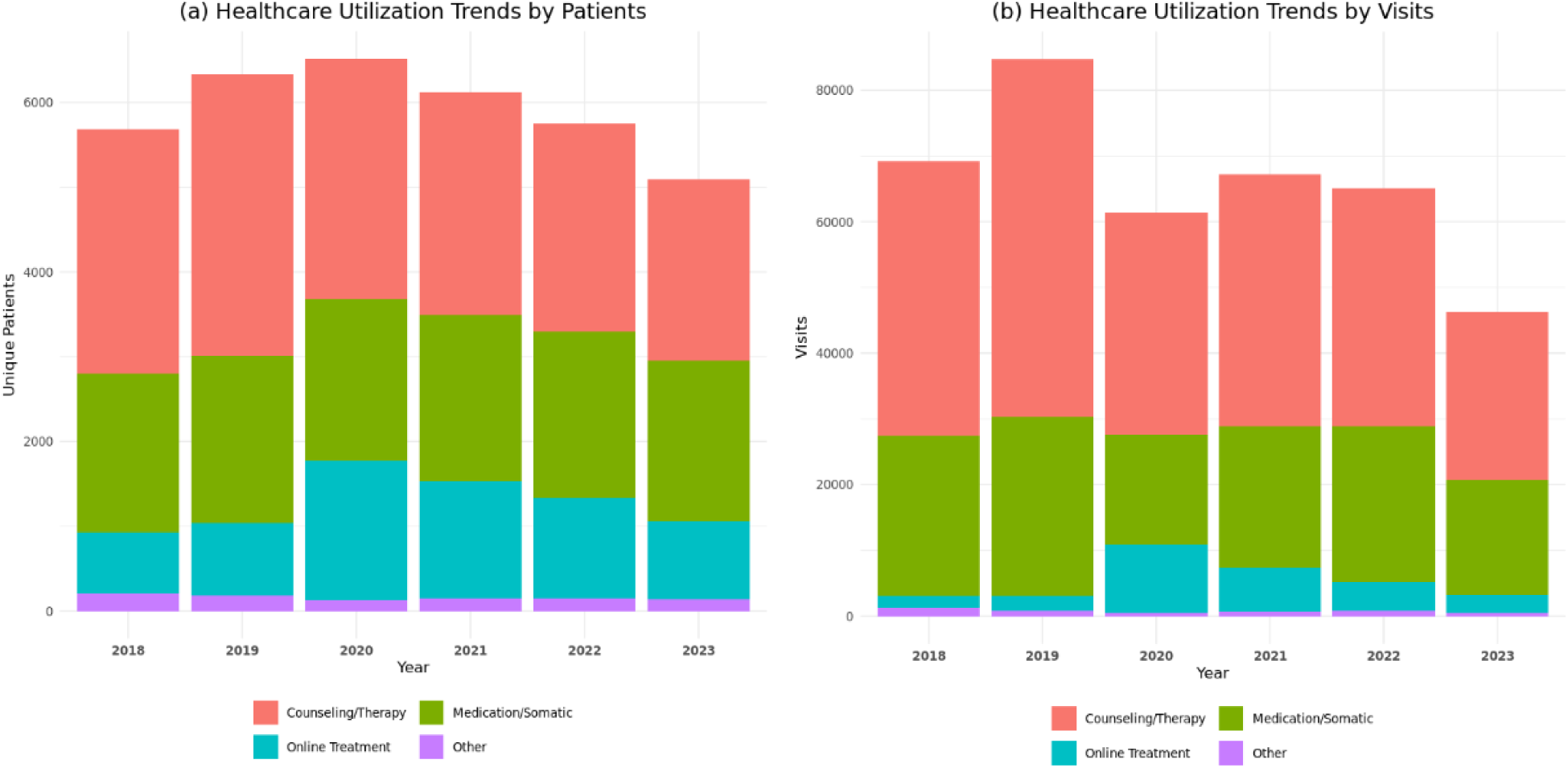
Healthcare utilization trends by healthcare service category, 2018–2023. *Healthcare utilization trends by healthcare service category among adults with co-occurring substance use and mental health disorders (COD), 2018–2023*. *(a) Trends by unique patients*. *(b) Trends by visits*. Counseling and therapy consistently accounted for the largest share of utilization across all years, while medication/somatic services remained relatively stable over time. Online healthcare peaked in 2020 during the COVID-19 pandemic before declining in subsequent years. The “Other” category represented the lowest-utilized services throughout the study period.

### Logistic Regression

Multivariable logistic regression identified several demographic and socioeconomic factors associated with healthcare receipt (*Table 2, Figure 4*).

**Figure 4.**
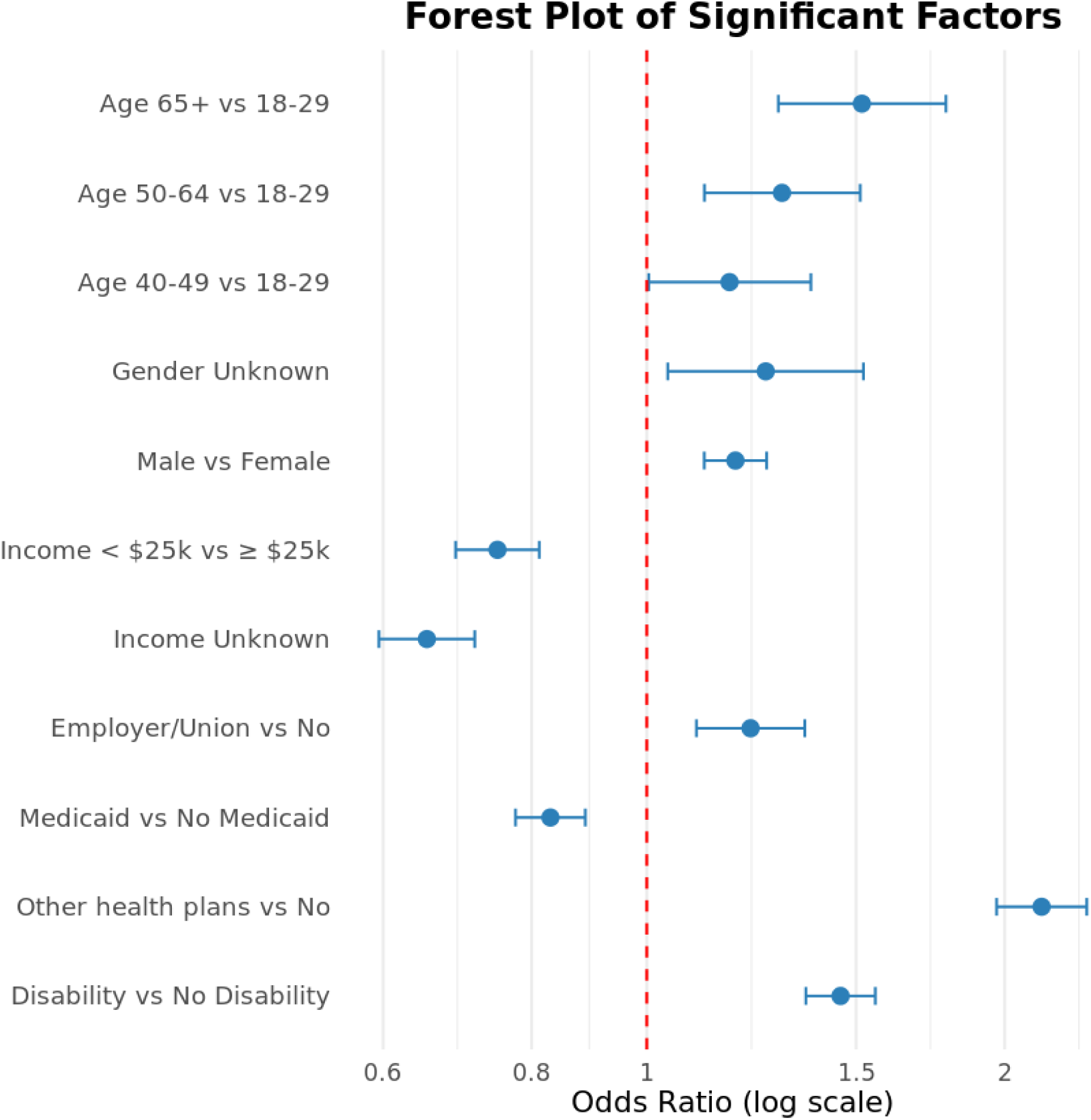
Forest plot of statistically significant sociodemographic and structural correlates of healthcare utilization among adults with co-occurring substance use and mental health disorders (COD). The figure displays adjusted odds ratios and 95% confidence intervals from the multivariable logistic regression model. Estimates are presented on a log scale, with the red dashed line indicating an odds ratio of 1.0 (no association).

**Table 2.**
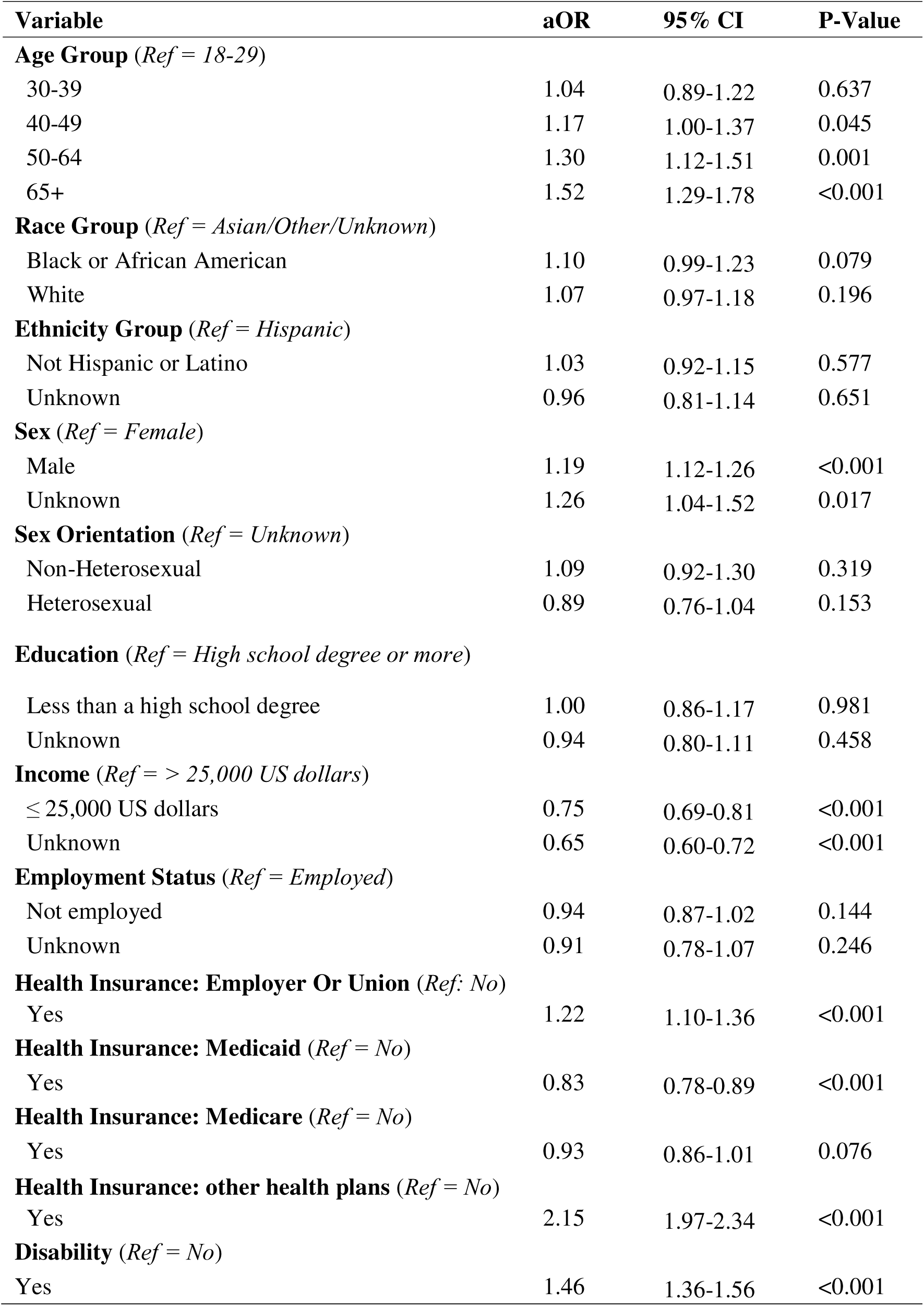
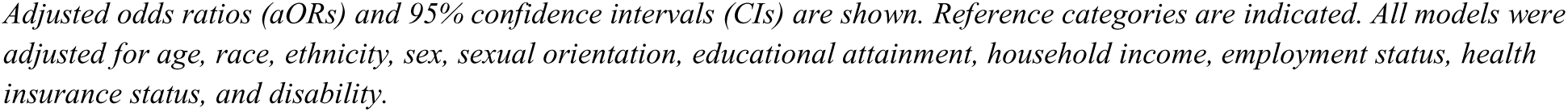
Multivariable logistic regression examining factors associated with healthcare utilization among adults with co-occurring substance use and mental health disorders.

| **Variable** | **aOR** | **95% CI** | **P-Value** |
| --- | --- | --- | --- |
| **Age Group** (*Ref = 18-29*) |  |  |  |
| 30-39 | 1.04 | 0.89-1.22 | 0.637 |
| 40-49 | 1.17 | 1.00-1.37 | 0.045 |
| 50-64 | 1.30 | 1.12-1.51 | 0.001 |
| 65+ | 1.52 | 1.29-1.78 | <0.001 |
| **Race Group** (*Ref = Asian/Other/Unknown*) |  |  |  |
| Black or African American | 1.10 | 0.99-1.23 | 0.079 |
| White | 1.07 | 0.97-1.18 | 0.196 |
| **Ethnicity Group** (*Ref = Hispanic*) |  |  |  |
| Not Hispanic or Latino | 1.03 | 0.92-1.15 | 0.577 |
| Unknown | 0.96 | 0.81-1.14 | 0.651 |
| **Sex** (*Ref = Female*) |  |  |  |
| Male | 1.19 | 1.12-1.26 | <0.001 |
| Unknown | 1.26 | 1.04-1.52 | 0.017 |
| **Sex Orientation** (*Ref = Unknown*) |  |  |  |
| Non-Heterosexual | 1.09 | 0.92-1.30 | 0.319 |
| Heterosexual | 0.89 | 0.76-1.04 | 0.153 |
| **Education** (*Ref = High school degree or more*) |  |  |  |
| Less than a high school degree | 1.00 | 0.86-1.17 | 0.981 |
| Unknown | 0.94 | 0.80-1.11 | 0.458 |
| **Income** (*Ref = > 25,000 US dollars*) |  |  |  |
| ≤ 25,000 US dollars | 0.75 | 0.69-0.81 | <0.001 |
| Unknown | 0.65 | 0.60-0.72 | <0.001 |
| **Employment Status** (*Ref = Employed*) |  |  |  |
| Not employed | 0.94 | 0.87-1.02 | 0.144 |
| Unknown | 0.91 | 0.78-1.07 | 0.246 |
| **Health Insurance: Employer Or Union** (*Ref: No*) |  |  |  |
| Yes | 1.22 | 1.10-1.36 | <0.001 |
| **Health Insurance: Medicaid** (*Ref = No*) |  |  |  |
| Yes | 0.83 | 0.78-0.89 | <0.001 |
| **Health Insurance: Medicare** (*Ref = No*) |  |  |  |
| Yes | 0.93 | 0.86-1.01 | 0.076 |
| **Health Insurance: other health plans** (*Ref = No*) |  |  |  |
| Yes | 2.15 | 1.97-2.34 | <0.001 |
| **Disability** (*Ref = No*) |  |  |  |
| Yes | 1.46 | 1.36-1.56 | <0.001 |

*Adjusted odds ratios (aORs) and 95% confidence intervals (CIs) are shown. Reference categories are indicated. All models were adjusted for age, race, ethnicity, sex, sexual orientation, educational attainment, household income, employment status, health insurance status, and disability.*

Sex and age were significant correlates of healthcare utilization. Compared with females, males had higher odds of receiving healthcare (aOR = 1.19, 95% CI: 1.12–1.26), as did participants with unknown sex designation (aOR = 1.26, 95% CI: 1.04–1.52). Relative to adults aged 18–29 years, healthcare receipt increased with age, with higher odds observed among those aged 40–49 years (aOR = 1.17, 95% CI: 1.00–1.37), 50–64 years (aOR = 1.30, 95% CI: 1.12–1.51), and ≥65 years (aOR = 1.52, 95% CI: 1.29–1.78). The 30–39 age group did not differ significantly from the youngest reference group.

Racial and ethnic differences were limited. Compared with the Asian/Other/Unknown reference group, Black or African American participants did not differ significantly in odds of healthcare receipt (aOR = 1.10, 95% CI: 0.99–1.23). No significant differences were observed for White participants or across ethnic groups. Sexual orientation and educational attainment were not significantly associated with healthcare receipt after adjustment.

Indicators of socioeconomic disadvantage were consistently associated with lower odds of healthcare utilization. Participants reporting annual household income ≤$25,000 (aOR = 0.75, 95% CI: 0.69–0.81) or unknown income (aOR = 0.65, 95% CI: 0.60–0.72) had substantially reduced odds of receiving healthcare. Employment status was not associated with healthcare receipt in adjusted analyses.

Health insurance coverage further differentiated patterns of service use. Medicaid coverage was associated with lower odds of healthcare receipt (aOR = 0.83, 95% CI: 0.78–0.89), while Medicare coverage was not significantly associated with utilization. In contrast, employer- or union-based insurance (aOR = 1.22, 95% CI: 1.10–1.36) and other health plans (aOR = 2.15, 95% CI: 1.97–2.34) were associated with higher odds of receiving healthcare. Disability status was also positively associated with healthcare receipt (aOR = 1.46, 95% CI: 1.36–1.56).

## Discussion

Using data from the nationally diverse All of Us Research Program (2018–2023), this study examined healthcare service utilization patterns among 19,423, adults with co-occurring substance use and mental health disorders (COD). Despite the complexity and elevated risk of COD, only 57.1% of participants received any healthcare during the study period. Counseling and therapy were the most used services, online healthcare showed a temporary surge during the COVID-19 pandemic, and medication/somatic service utilization remained stable throughout. Healthcare service utilization varied significantly by sociodemographic and structural determinants, including age, gender, income, insurance type, and disability status, highlighting persistent inequities in access and engagement.

The 57.1 % healthcare utilization proportion of this study cohort remains below the national estimate of 62.4 % among adults with COD, as reported by the 2023 National Survey on Drug Use and Health (Substance Abuse and Mental Health Services Administration, 2024b) and is consistent with prior national analyses documenting low and fragmented healthcare engagement in this population (Jones & McCance-Katz, 2019). Differences in measurement methodology may partly explain this gap: national estimates rely on self-reported service use, whereas the present study draws on EHR data that capture only documented encounters within participating systems. Individuals who accessed community-based or informal care outside of clinical settings may therefore not have been represented. Moreover, the All of Us cohort intentionally engages individuals historically under-represented in biomedical research, including individuals those from lower socioeconomic backgrounds (All of Us Research Program Investigators, 2019). Recent phenomic analyses of All of Us participants have also shown that the cohort differs systematically from the general U.S. population, indicating that findings from All of Us may not be fully generalizable to national patterns of COD healthcare utilization (Zeng et al., 2024).

Collectively, these findings emphasize persistent limitations in service engagement for people with COD and the need for stronger coordination across systems of care.

Counseling and therapy services emerged as the dominant healthcare approach, accounting for the largest share of utilization. This pattern aligns with prior work emphasizing the centrality of psychotherapy in behavioral health care (Flynn & Brown, 2008; Johansen & Sicker, 2022). The broad applicability of psychotherapy billing codes and the widespread availability of counseling services likely contribute to their prominence. Psychosocial counseling interventions remain the most empirically supported first-line healthcare services for both MHD and SUD, making them a common initial modality of care for individuals with COD (Azad et al., 2022; Carroll & Kiluk, 2017).

Beyond counseling, medication and somatic service utilization was stable throughout the study period, highlighting the continued importance of pharmacological interventions for managing symptoms of severe mental illness and substance use disorders (Iqbal et al., 2019; Simpson et al., 2021). More intensive services, such as detoxification, residential programs, and crisis intervention, were rarely utilized. This underutilization reflects persistent structural barriers, including cost, availability, and fragmentation between mental health and addiction systems (Foster et al., 2010; Priester et al., 2016). The separation of funding streams and licensing requirements may further inhibit access to integrated, high-intensity care (Priester et al., 2016).

High costs and limited geographic availability of residential programs also disproportionately burden low-income and rural populations, widening existing disparities (Lynch et al., 2024).

There were substantial shifts in temporal trends during the COVID-19 pandemic. Counseling and therapy services peaked in 2019 and subsequently declined, likely reflecting disruptions in in-person care delivery and the broader psychosocial strain of the pandemic. This trajectory mirrors national evidence documenting increased psychological distress and substance use during this period, which reshaped patterns of care-seeking (Czeisler et al., 2020). In contrast, online healthcare services surged in 2020 amid lockdowns and the rapid expansion of telehealth infrastructure, reflecting a temporary shift toward virtual modalities (Uscher-Pines et al., 2022). Despite this rise, online healthcare declined in subsequent years, suggesting that telehealth engagement may be difficult to sustain among individuals with COD. Barriers such as limited digital access, provider capacity, and challenges in delivering integrated dual-diagnosis care virtually likely contributed to this decline (Kisicki et al., 2022; Pham et al., 2025).

Findings revealed associations between several demographic and structural factors and healthcare utilization. Older adults were more likely to receive care, consistent with evidence showing lower utilization among younger populations despite their higher prevalence of COD (Dhinsa et al., 2023; Substance Abuse and Mental Health Services Administration, 2024b). This age pattern may also reflect differences in insurance coverage, because older adults are more likely to have stable health insurance, including Medicare, which increases access to behavioral health services compared with younger adults who experience higher rates of uninsurance (Bunch & Ketema, 2025). These differences in coverage and access may compound the challenges younger adults already face. Barriers for younger adults may include stigma, limited age-appropriate services, and insufficient outreach (James et al., 2024; Sterling et al., 2010).

Gender disparities were also evident: male participants and those with an unspecified gender designation had higher odds of receiving healthcare than female participants, contrasting with national trends of greater engagement among women (Dhinsa et al., 2023). Barriers such as childcare responsibilities, trauma histories, and stigma may restrict service use among women with COD (Harris et al., 2022; Rosen et al., 2004).

Socioeconomic status exerted a substantial influence on service utilization. Lower income was associated with reduced healthcare utilization, reinforcing the role of financial and systemic barriers in limiting access. This pattern aligns with evidence showing that low-income individuals face persistent cost-related and geographic obstacles to both mental health and substance use healthcare (Edlund et al., 2009; Kim & Cardemil, 2012). While some research suggests higher SUD healthcare rates among low-income groups due to targeted outreach, these dynamics may differ in COD populations, where access to integrated and continuous care remains constrained (Dhinsa et al., 2023).

Insurance coverage further shaped access. Participants with employer-sponsored or private insurance were significantly more likely to receive healthcare, whereas Medicaid recipients were less likely. This discrepancy may reflect state-level variation in Medicaid behavioral-health benefits, provider shortages, and administrative burdens that limit comprehensive coverage.

Differences in reimbursement rates and coverage for residential or intensive programs further restrict service availability, particularly for individuals with COD (Dickson-Gomez et al., 2022; Foster et al., 2010; Priester et al., 2016). Addressing these inequities will require policies that standardize and expand Medicaid behavioral-health benefits to ensure consistent, integrated care across states.

Disability status was positively associated with healthcare utilization, suggesting that individuals with more complex health needs may be more likely to access services. This finding aligns with prior evidence showing that adults with disabilities experience higher prevalence of MHD and SUD and have greater contact with healthcare systems, resulting in increased service utilization (Reif et al., 2022). However, despite higher engagement, many still encounter barriers such as accessibility challenges, provider stigma, and financial constraints, which may limit continuity and adequacy of care (Xie et al., 2022).

### Limitations

This study has several limitations. Although the primary analyses examined healthcare utilization at the individual level, descriptive summaries of service trends were based on encounter counts, which may overrepresent individuals with more frequent visits. In addition, the use of EHR data limited the ability to distinguish whether services were provided specifically for substance use disorders, mental health disorders, or integrated co-occurring disorder care, because many CPT and ICD codes are not diagnosis-specific.

Some CPT- and HCPCS-defined procedure codes were not available in the All of Us database. Consequently, certain healthcare-related encounters may not have been captured in this analysis. This incomplete availability of procedure code data may have resulted in incomplete measurement of healthcare utilization, particularly if the absence of specific codes was not random. Finally, because the All of Us cohort only consists of individuals engaged in the participating healthcare systems, findings may not generalize to individuals receiving community-based, informal, or non-system-affiliated care that have not been included in the All of Us Program.

### Conclusions

Despite these limitations, the findings offer important insights into healthcare utilization among adults with COD. The persistent underutilization of care emphasizes the importance of strengthening integrated models that address both mental health and substance-use conditions concurrently. Disparities observed across sociodemographic and structural factors further reflect systemic inequities in access and engagement that warrant equity-focused policy and system-level responses. Expanding and standardizing Medicaid behavioral-health benefits, improving provider availability, and investing in culturally responsive services remain important priorities for promoting more consistent and integrated care. The temporary rise and subsequent decline in telehealth utilization also represent a missed opportunity to sustain care modalities capable of reducing structural barriers such as transportation challenges and geographic isolation. Building sustainable, inclusive digital-care infrastructure will be essential to improving long-term engagement for individuals with complex behavioral-health needs. Future research should examine how system-level reforms and digital innovations can be integrated to build a more equitable, sustainable continuum of behavioral-health care for individuals with COD.

## Funding

This study was supported by the NIH/NIAID grant (#R01AI174892) and NIH/NIMH grant (#R01MH127961-02S1).

## Role of the Funding Source

The funding sources had no role in the study design; analysis, or interpretation of data; writing of the manuscript; or the decision to submit the manuscript for publication.

## Author Contributions

ASI conceptualized the study and contributed to the original draft writing, review, and editing. MW performed the formal analysis and contributed to the original draft writing, review, and editing. ZB contributed to the original draft writing, review, and editing. XL contributed to study conceptualization and provided critical review and editing of the manuscript for important intellectual content. SQ contributed to study conceptualization, provided critical review and editing of the manuscript for important intellectual content, and supervised the overall conduct of the study. All authors read and approved the final version of the manuscript.

## Data Availability Statement

The data used in this study are available through the National Institutes of Health All of Us Research Program.

## Competing Interests

The authors declare no potential conflicts of interest with respect to the authorship, research, or publication of this article

## Supporting information

CPT code

## Notes

### Competing Interest Statement

The authors have declared no competing interest.

### Funding Statement

This study was supported by the NIH/NIAID grant and NIH/NIMH grant.

### Author Declarations

The study used ONLY openly available human data that were originally located at: https://workbench.researchallofus.org/login?_gl=1*row0yf*_ga*NDgxODQ3MjA3LjE3NjE3NTY5NDQ.*_ga_MQVR5DG2C4*czE3NjMxNTQ5NDYkbzMkZzAkdDE3NjMxNTQ5NDYkajYwJGwwJGgzOTUwNzI5NjU.

