## Supplementary material for "Healthcare utilization among adults with co-occurring substance use and mental health disorders (2018-2023): A study based on All of Us program": CPT code

Supplementary Table 1. ICD-10-CM codes used to define substance use disorders (SUD) and mental health disorders (MHD)

| ICD 10 Code | Phecode description |
| --- | --- |
| F10 | Alcohol-related disorders/ Alcoholism |
| F11 | Opioid-related disorders |
| F12 | Cannabis-related disorders |
| F13 | Sedative-related disorders |
| F14 | Cocaine-related disorders |
| F15 | Other stimulant-related disorders |
| F16 | Hallucinogen-related disorders |
| F17 | Tobacco use disorder |
| F18 | Inhalant-related disorders |
| F19 | Other psychoactive substances |
| F20 | Schizophrenia and other psychotic disorders |
| F25 | Schizoaffective disorders |
| F31 | Bipolar disorder |
| F32 | Major depressive disorder, single episode |
| F33 | Major depressive disorder, recurrent |
| F34 | Dysthymic disorder/ Cyclothymic disorder/ Other persistent mood [affective] disorders |
| F39 | Unspecified mood [affective] disorder |
| F40 | Specified phobia and anxiety disorders |
| F41 | Other anxiety disorders |
| F42 | Obsessive-compulsive disorder |
| F43 | Post-traumatic stress disorder (PTSD) |
| F50 | Eating disorder |
| F51 | Sleep disorders |
| F60 | Personality disorders |
| F90 | Attention-deficit hyperactivity disorder |
| F91 | Conduct disorders |

Supplementary Table 2. CPT and HCPCS codes used to identify substance use disorder (SUD) and mental health disorder (MHD) healthcare services

|  | SUD CPT Codes | SUD HCPCS Codes | MHD CPT Codes | MHD HCPCS Codes |
| --- | --- | --- | --- | --- |
| Counseling, Therapy, and Case Management | 90791, 90792,90832, 90834, 90837, 90846, 90847, 90849, 90853, | H0004, H0005, H0006, H2027, H2032, G0396, G0397, G0443, G2011, T1006, T1007 | 90887, 90889, 90845, 99492, 99493, 99494, 90888*, 99848* | G2087, G2086, G2088*, H0036* |
| Crisis Intervention & Acute Care | 90839, 90840, | H0007, H2011, H2012, H2013* |  |  |
| Detoxification Services |  | H0008, H0009, H0010, H0011, H0012, H0013, H0014 |  |  |
| Intensive Outpatient & Treatment Programs |  | H0015, H2035, H2036, H2001* |  |  |
| Residential & Halfway House Programs |  | H0017, H0018, H0019, H2034* |  |  |
| Medication & Medical/Somatic Services | 99202, 99203, 99204, 99205, 99211, 99212, 99213, 99214, 99215, 99408, 99409, 99206*, 99207*, 99208*, 99209*, 99210* | H0016, H0020, G2067, G2068, H2010*, G2213* | 90863 |  |
| Rehabilitation & Supportive Services |  | H2017, H2018, H2019, H2015*, H2016*, H2020*, H2021*, H2022*, H2030*, H2031*, H2038*, T1012* |  |  |
| Specialized or other treatment services |  | H0022, H2040*, H2041* | 90899 |  |
| Online treatment | 99441, 99442, 99443, 99421, 99422, 99423, | G2010 |  |  |

*Codes marked with an asterisk (*) were not observed in the All of Us dataset but were included to document the full set of CPT and HCPCS codes reviewed for treatment identification.*
